# The protective association between statins use and adverse outcomes among COVID-19 patients: a systematic review and meta-analysis

**DOI:** 10.1101/2021.02.08.21251070

**Authors:** Ronald Chow, James Im, Nicholas Chiu, Leonard Chiu, Rahul Aggarwal, Jihui Lee, Young-Geun Choi, Elizabeth Horn Prsic, Hyun Joon Shin

## Abstract

**Introduction:** Statins may reduce a cytokine storm, which has been hypothesized as a possible mechanism of severe COVID-19 pneumonia. The aim of this study was to conduct a systematic review and meta-analysis to report on adverse outcomes among COVID-19 patients by statin usage.

**Methods:** Literatures were searched from January 2019 to December 2020 to identify studies that reported the association between statin usage and adverse outcomes, including mortality, ICU admissions, and mechanical ventilation. Studies were meta-analyzed for mortality by the subgroups of ICU status and statin usage before and after COVID-19 hospitalization. Studies reporting an odds ratio (OR) and hazard ratio (HR) were analyzed separately.

**Results:** Thirteen cohorts, reporting on 110,078 patients, were included in this meta-analysis. Individuals who used statins before their COVID-19 hospitalization showed a similar risk of mortality, compared to those who did not use statins (HR 0.80, 95% CI: 0.50, 1.28; OR 0.62, 95% CI: 0.38, 1.03). Patients who were administered statins after their COVID-19 diagnosis were at a lower risk of mortality (HR 0.53, 95% CI: 0.46, 0.61; OR 0.57, 95% CI: 0.43, 0.75). The use of statins did not reduce the mortality of COVID-19 patients admitted to the ICU (OR 0.65; 95% CI: 0.26, 1.64). Among non-ICU patients, statin users were at a lower risk of mortality relative to non-statin users (HR 0.53, 95% CI: 0.46, 0.62; OR 0.64, 95% CI: 0.46, 0.88).

**Conclusion:** Patients administered statins after COVID-19 diagnosis or non-ICU admitted patients were at lower risk of mortality relative to non-statin users.

## INTRODUCTION

In December 2019, the first outbreak of a novel human coronavirus infection named severe acute respiratory syndrome-related coronavirus 2 (SARS-CoV-2) was reported in Wuhan, China^1^. The SARS-CoV-2 then rapidly spread across the globe, manifesting a high incidence of COVID-19 disease; on March 12, 2020, the World Health Organization declared COVID-19 as a pandemic^2^.

The pathophysiology that underlies the severe COVID-19 pneumonia is an overproduction of early response proinflammatory cytokines, namely tumour necrosis factor (TNF), IL-6 and IL-1β^3^. If left unattended, this cytokine storm could place COVID-19 patients at a higher risk of vascular hyperpermeability, multiorgan failure, and death^3^. Moreover, cholesterol has been implicated to have a possible role in an increased risk of infection in the elderly patients, wherein higher tissue cholesterol has been shown to increase the endocytic entry of SARS-CoV-2. Statins, with its mechanism of inhibiting 3-hydroxy-3-methyl-glutaryl-CoA (HMG-CoA) reductase inside cells, may reduce such a cytokine storm, as it stabilizes the protein myeloid differentiation primary response 88 (MYD88) at normal levels, and also up-regulates angiotensin-converting enzyme 2 (ACE2), which is reported to be downregulated by SARS-CoV-2 and decrease infiltration of SARS-CoV-2 into the cells^4-6^.

Several studies have reported on the use of statins amongst COVID-19 patients with differing conclusions. Zhang *et al*^7^ reported that the use of statin was associated with reduced mortality, while Cariou *et al*^8^ reported an opposite association with an increased mortality. Other studies such as De Spiegeleer *et al*^9^and Song *et al*^10^ reported no difference in mortality between statin and non-statin users. Further studies, or meta-analyses are sorely needed to pool study results and increase statistical power, to achieve a better understanding of the potential effect of statins.

Meta-analyses by Hariyanto *et al*^11^ and Kow *et al*^12^ have studied the association between statin use and mortality among COVID-19 patients. However, while they analyzed several studies on the association between statins and adverse outcomes on COVID-19 patients, neither meta-analyses comprehensively reported on all studies conducted to-date of their publication. Further, neither paper distinguished studies on the circumstances around statin use; specifically, whether included studies in their meta-analysis reported on patients who used statin prior to hospitalization and statins after hospitalization.

The aim of this study was to conduct a systematic review and meta-analysis to report on adverse outcomes among COVID-19 patients by statin usage.

## METHODS

### Search Strategy

Ovid MEDLINE, Embase and the Cochrane Central Register of Controlled Trials were searched from January 2019 to December 2020, for English-language publications. The search strategy is presented in Appendix 1.

### Study Screening

Studies underwent level 1 title and abstract screening to identify articles that reported on the use of statins among COVID-19 patients. Relevant articles from level 1 screening subsequently underwent level 2 full-text screening. This step identified studies that reported on a clinical database for which adverse outcomes were reported. Reference lists of articles undergoing level 2 full-text screening were also assessed (i.e. backward reference searching), to identify other potentially relevant studies.

All screening was done independently by two reviewers (RC, JI). Where there were disagreements, a discussion occurred and consensus was achieved to determine whether to include or exclude an article. Where consensus could not be achieved, a third author (NC) was consulted for their input and final decision.

### Quantitative Synthesis

Articles identified for inclusion after level 2 full-text screening were assessed for quantitative synthesis. Articles were included if they reported an adjusted relative risk measure on mortality, ICU admissions or mechanical ventilation of patients who were statin users compared to those who were not statin users (comparison group).

For studies included at this stage, demographics such as sample size, study design, patient population, central measures of tendency for age, percentage male, and percentage of statin users were noted. We noted whether studies reported exclusively on ICU patients at the time of enrollment. Additionally, we assessed if statin use was defined as prior to or after hospitalization.

In corollary to study screening, quantitative synthesis was conducted independently by two reviewers (RC, JI), and a third reviewer (NC) was consulted if consensus was not achieved.

### Statistical Analyses

Meta-analysis was conducted for mortality by the subgroups of ICU status (ICU Patients, and Non-ICU Patients) and statin usage (Statin Use Before and After COVID-19 Hospitalization). Studies employing similar relative risk ratios were analyzed together to generate a summary risk estimate. That is, studies reporting an odds ratio (OR) were aggregated to produce a summary odds ratio and studies reporting a hazard ratio (HR) were analyzed together. Corresponding 95% confidence intervals (CI) for an OR or a HR are reported as well. When there was heterogeneity of I^2^ of equal to or greater than 50%, a random-effects DerSimonian-Laird analysis model was applied; otherwise, a fixed-effects inverse-variance analysis model was used. A *p*-value of less than 0.05 was considered statistically significant.

A funnel plot and Egger’s test were used to assess the publication bias for the endpoint of mortality. A *p*-value of greater than 0.05 indicated a lack of asymmetry of the funnel plot, and thereby no significant concern for publication bias. All analyses were conducted using Stata 16.

## RESULTS

A total of 482 records underwent level 1 screening, of which 474 were identified through database search and 8 identified from backward reference searching. After removing duplicates, 346 records underwent level 1 screening; 49 articles subsequently underwent level 2 screening. Twenty-one studies were assessed for quantitative synthesis, of which 13 studies^7-10,13-22^, presenting on 14 cohorts, reported relative risk ratios adjusted for potential confounders and were included in this systematic review and meta-analysis (Appendix 2).

Study demographics are presented in Table 1. All studies were observational in design, with either a retrospective cohort study design or a case-control study design. Israel *et al*^16^ split their database into two cohorts and reported two relative risk ratios for the outcome of mortality. Two studies, Grasselli *et al*^14^ and Rodriguez-Nava *et al*^20^, reported on ICU patients; the other 11^7-10,13,15-19,21,22^ reported on non-ICU patients. Six studies^7,10,17,18,20,21^ defined a statin user as one who used statins after a COVID-19 hospitalization; seven studies^8,9,13-16,19,22^ defined a statin user as one who used statins before a COVID-19 hospitalization.

**Table 1.**
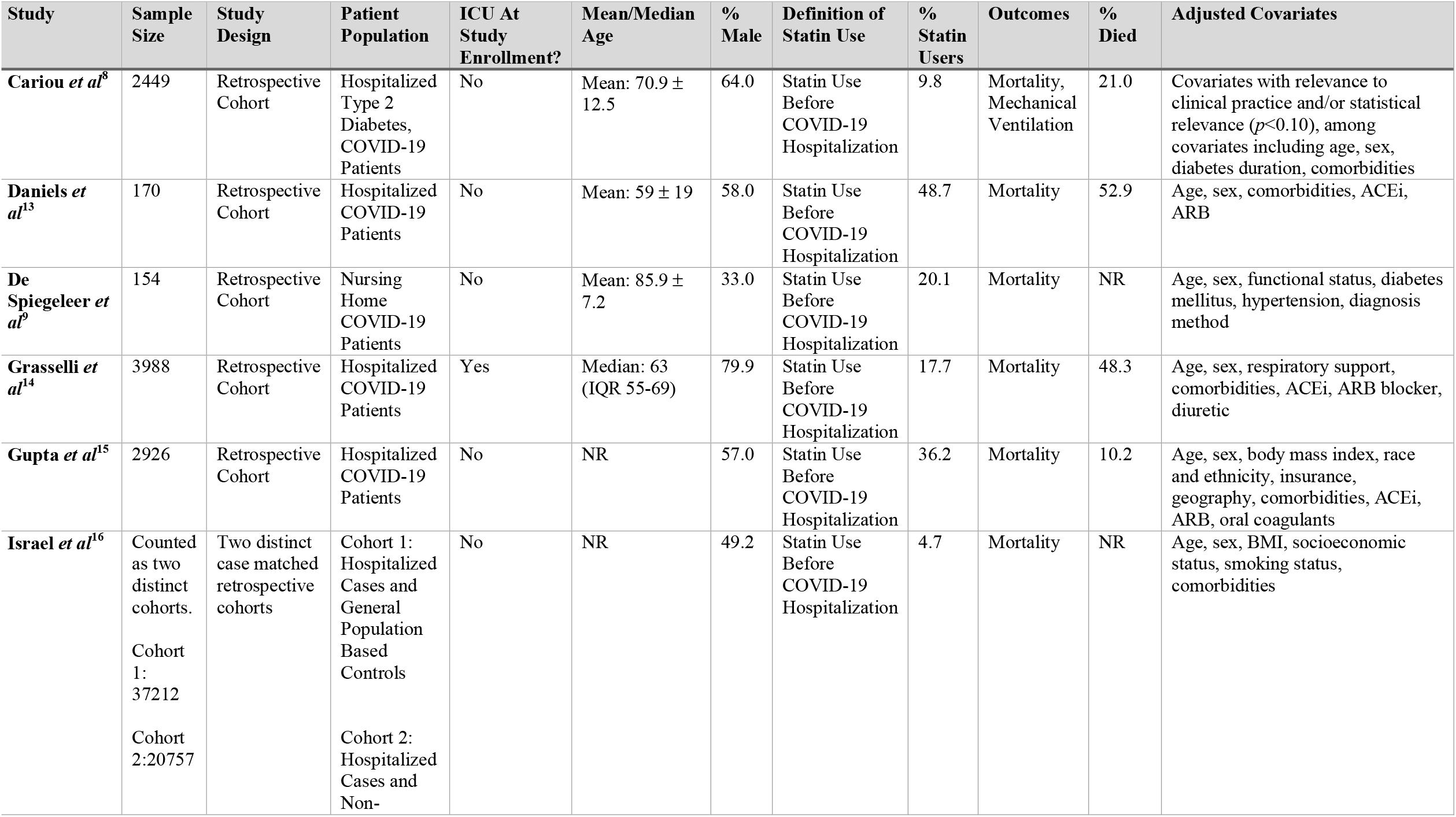
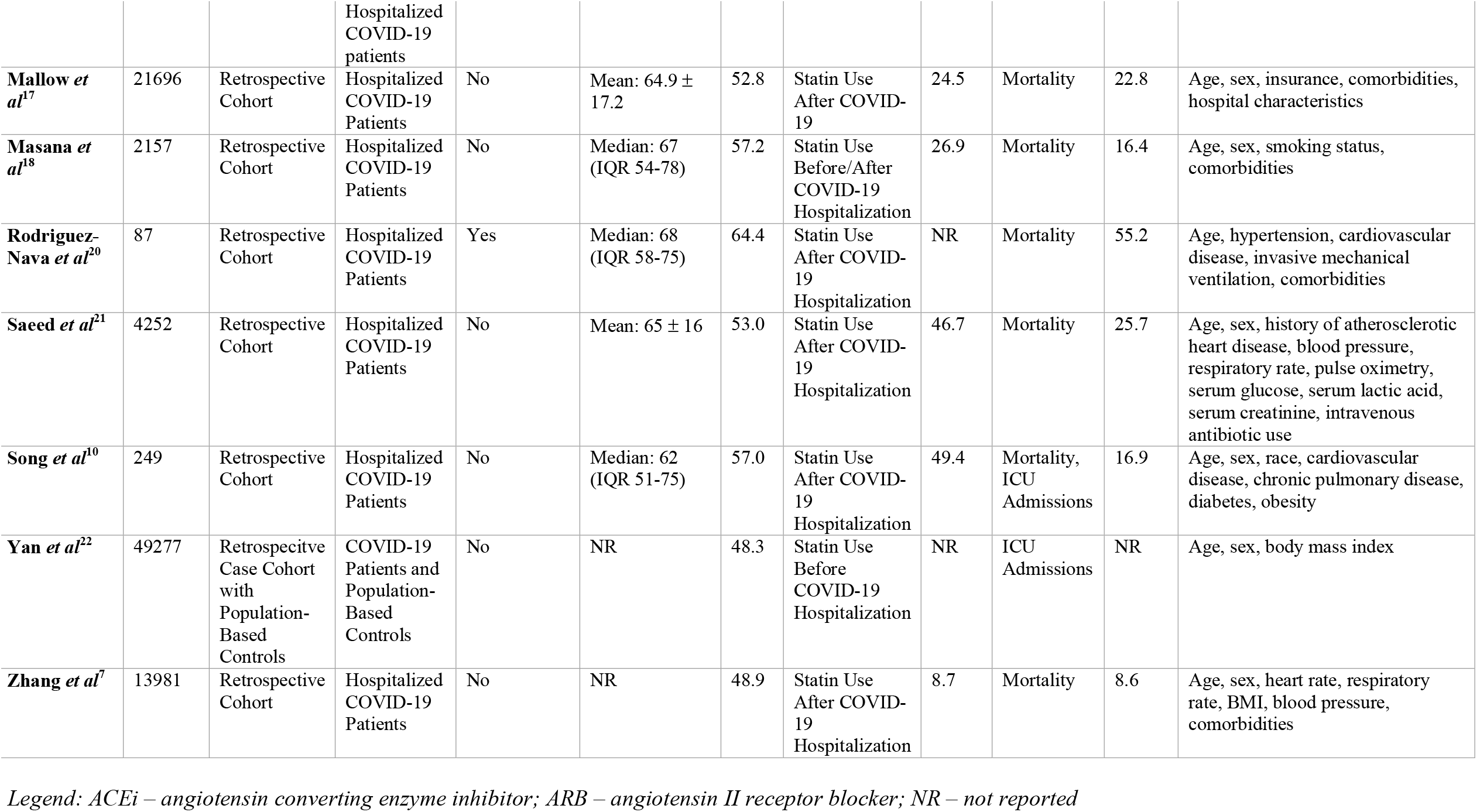
Study Demographics

Visual inspection of the funnel plot and Egger’s test (*p* = 0.086) suggests no significant concern for publication bias (Appendix 3).

### Mortality

Twelve studies compared mortality between statin users and non-users. Individuals who had used statins prior to COVID-19 hospitalization had a similar risk of mortality to those who did not use statins – HR of 0.80 (95% CI: 0.50, 1.28) and OR of 0.62 (95% CI: 0.38, 1.03). COVID-19 patients who had statins administered after their COVID-19 hospitalization had a lower risk of mortality compared to those who did not receive statins – HR of 0.53 (95% CI: 0.46, 0.61) and OR of 0.57 (95% CI: 0.43, 0.75) (Figure 1a).

**Figure 1.**
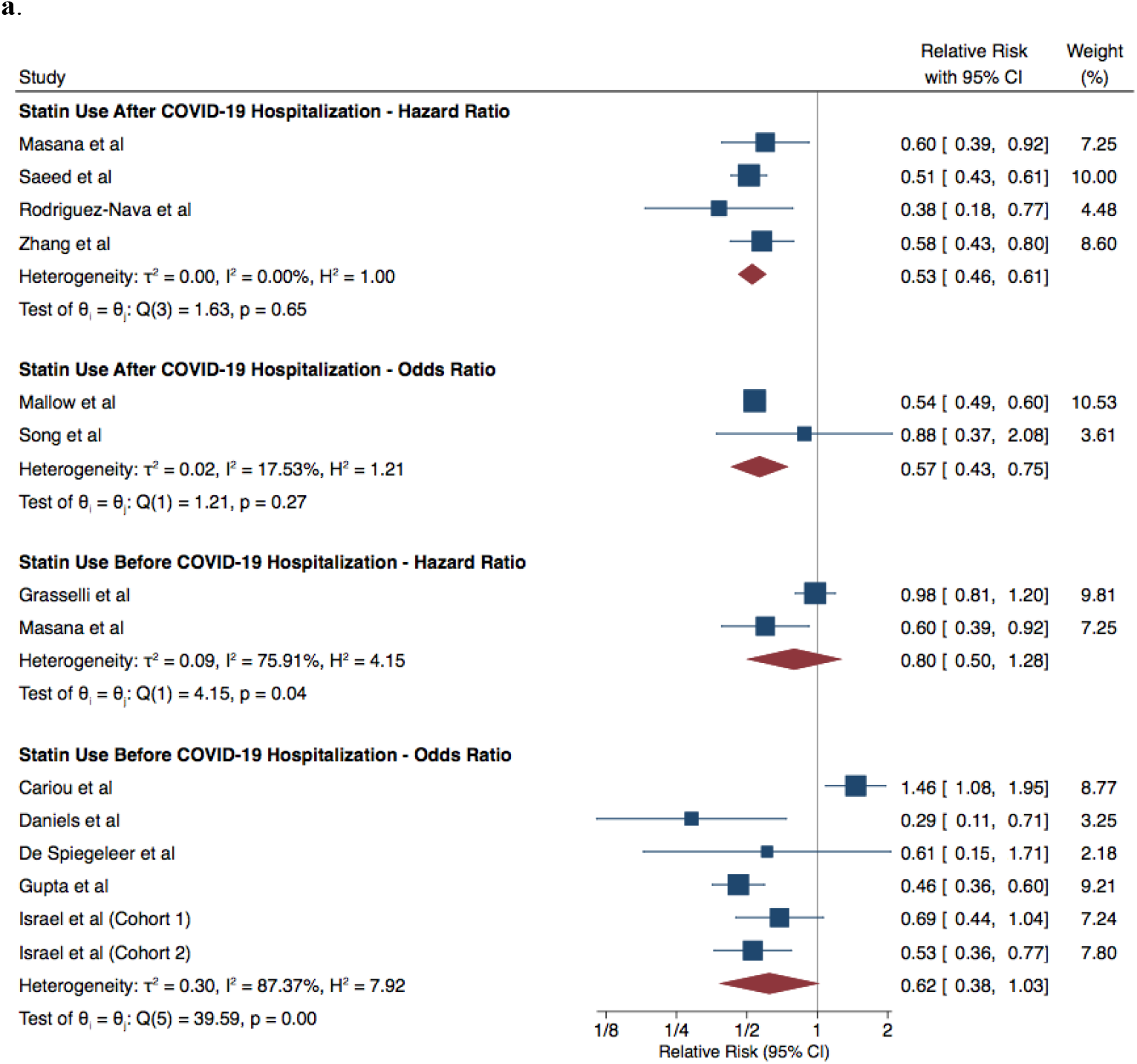
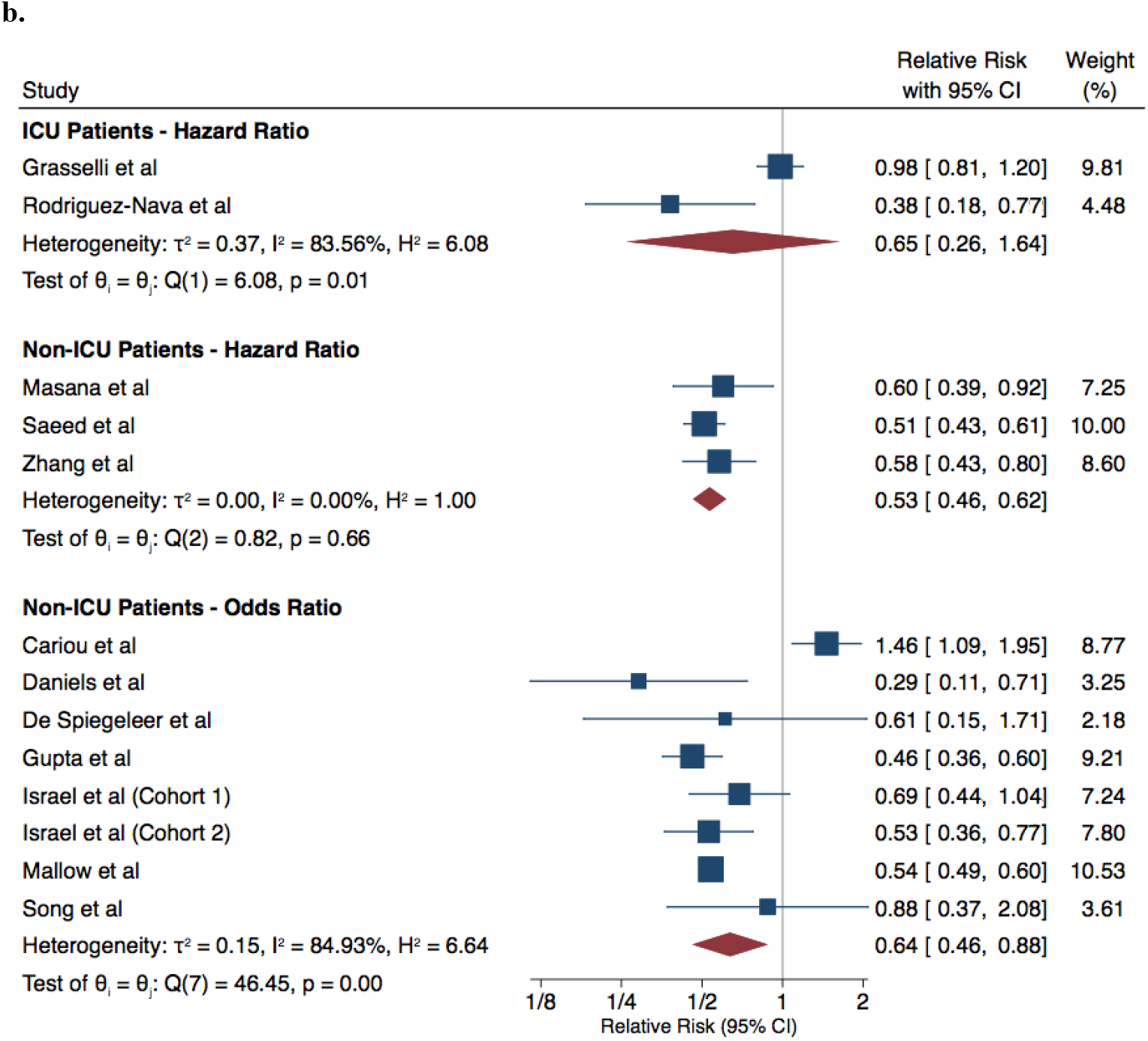
COVID-19 Patients Who Used Statins vs Did Not Use Statins – Mortality Analysis **1a** By Statin Usage **1b** By ICU/Non-ICU Patients

COVID-19 patients who were using statins and admitted to the ICU did not show a lower risk of mortality, compared to non-statin users – OR of 0.65 (95% CI: 0.26, 1.64). Among non-ICU patients, statin users were at a lower risk of mortality relative to non-statin users – HR of 0.53 (95% CI: 0.46, 0.62) and OR of 0.64 (95% CI: 0.46, 0.88) (Figure 1b).

### ICU Admissions

Two studies reported on ICU admissions by patients who did and did not use statins. The risk of ICU admissions was similar between patients who used statins before COVID-19 hospitalization and who did not use statins, according to Yan *et al* – OR of 1.78 (95% CI: 0.54, 5.13). It was also similar between patients who used and did not use statins after COVID-19 hospitalization, according to Song *et al* – OR of 0.90 (95% CI: 0.49, 1.67).

### Mechanical Ventilation

Cariou *et al* reported the odds of mechanical ventilation to be 0.99 (95% CI: 0.69, 1.41), when comparing statin to non-statin users.

## DISCUSSION

This is the first rigorously conducted systematic review and meta-analysis investigating the association between statin use and adverse outcomes among COVID-19 patients, meeting the methodological guidance for a high quality systematic review and meta-analysis as recommended by the MOOSE group^23^. With thirteen cohorts reporting on 110,078 patients in total, this meta-analysis has much greater statistical power than the meta-analyses of Hariyanto *et al*^11^ and Kow *et al*^12^. The analysis by Hariyanto *et al*^11^ reported on 9 studies and an amalgamated sample size of 3,263 while Kow *et al*^12^ reported on 4 studies and a total sample size of 67,333.

Furthermore, this review employs different statistical methodologies than the prior reviews. While Kow *et al*^12^ jointly meta-analyzed study outcomes of mortality and severity together, we separately analysed the outcomes of mortality and ICU admissions. This review also differs from the Hariyanto *et al*^11^ paper in that we meta-analyzed adjusted relative risk ratios, while they meta-analyzed unadjusted event data. As all the studies employ an observational study design, covariate distributions of statin user and non-user groups may not necessarily be similar as one would expect in a randomized controlled trial. A meta-analysis of adjusted relative risk ratios excluding unadjusted results is necessary.

The results suggest that individuals with and without history of statin use before their COVID-19 hospitalization experience a similar risk of mortality, ICU admissions, and use of mechanical ventilation. However, we urge caution around this simple dichotomy of studies and the corresponding interpretation. Depending on the time sequence of events, we hypothesize there are four possible scenarios of statin use within these included studies (Figure 2):

**Figure 2.**
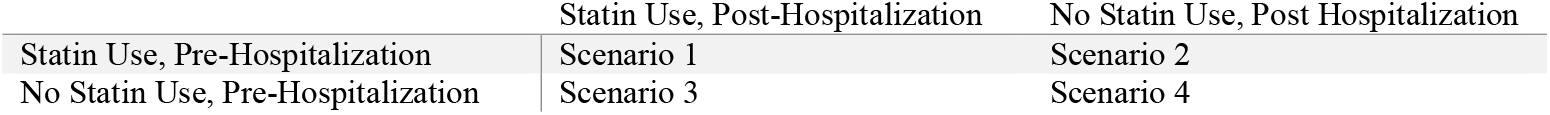
Potential Patient Scenarios

1. Patients who used statins prior to COVID-19 hospitalization and who continue to use statins after hospitalization
2. Patients who used statins prior to COVID-19 hospitalizations and who cease to use of statins after hospitalization
3. Patients who have no history of statin use prior to COVID-19 hospitalization and start using statins after hospitalization
4. Patients who have no history of statin use prior to COVID-19 hospitalization and do not receive statins after hospitalization

As statins are not currently a recommended medication for COVID-19 treatment and there is no randomized controlled trial in this setting, one could assume that Scenario 3 is unlikely to happen given its intent to solely treat COVID-19 with statin.

In the event that Scenario 3 did occur, patients in Scenario 3 who only used statins after COVID-19 hospitalization may have acute cardiovascular indications for statin use after hospitalization, such as those who may develop acute myocardial injury, acute coronary syndrome or stroke. Our analysis by statin use before COVID-19 hospitalization therefore compares patients who used statins prior to COVID-19 hospitalization who may or may not continue using statins (Scenarios 1 and 2) after hospitalization, to patients who did not use statins prior to COVID-19 hospitalization who may or may not start statins after hospitalization (Scenarios 3 and 4). The data reporting on non-statin users prior to COVID-19 hospitalization would therefore include more patients with poorer outcomes (Scenario 3), and result in an overestimation of the possible protective association between statin use before hospitalization and adverse outcomes.

Along a similar vein, the possible protective association of statins prior to COVID-19 hospitalization and adverse outcomes may also be underestimated. Masana *et al*^*18*^ reported further on Scenario 2, in which 42.2% of enrolled patients who used statins before COVID-19 hospitalization but discontinued statins during hospitalization. While the pre-hospitalization analysis would not be affected, the hidden assumption of pre-hospitalization statin use reflecting, or being used as proxy for, post-hospitalization statin use would be violated; the association between statin use before COVID-19 hospitalization and adverse outcomes will underestimate the association between statin after COVID-19 hospitalization and adverse outcomes. Hence, by comparing studies based on whether they use statins before COVID-19 hospitalization as a proxy for statin use after COVID-19 may result in a significant misclassification.

This review also reports that patients who used statins after their COVID-19 hospitalization were at a lower risk of mortality and this protective association between statins use and mortality may be underestimated. Of the four aforementioned scenarios, our comparison of statin use after COVID-19 hospitalization may be summarized as a comparison of patients who used statins after COVID-19 hospitalization regardless of prior usage (Scenarios 1 and 3), relative to patients who did not use statins after COVID-19 hospitalization regardless of prior usage (Scenarios 2 and 4). Again, in the event that Scenario 3 manifested and thereby the arm of patients who used statins after COVID-19 hospitalization experience poorer outcomes, the protective association between statins use after COVID-19 hospitalization and mortality may be underestimated.

Masana *et al* also reported that patients who continued on statins had significantly better outcomes, compare to patients who were not on statins before COVID-19 (*p*=0.045). In fact, our results suggest that statin usage after COVID-19 hospitalization may have a protective association for mortality, and our analysis supports the recommendation by UpToDate® for “continuing statins in hospitalized patients with COVID-19 who are already taking them”^24^. However, prospective randomized controlled trials should further investigate whether statins may be an effective therapy for COVID-19 patients.

Our analysis also suggests that the risk of mortality among statin users compared to non-statin users differs, when focusing on subsets of ICU and non-ICU patients. Non-ICU patients seemed to experience benefit from statin use, while this beneficial association was not seen among patients admitted to the ICU. It is possible that statin has a more beneficial effect to decrease inflammation, early in the stage of illness and among less sick patients with COVID-19. However, there is a paucity of data with respect to ICU patients. Further investigation is required among ICU patients, given this limited statistical power to find association between statin and mortality, among ICU patients.

This study was not without limitations. Intrinsic to meta-analysis study designs, the strength of meta-analysis conclusions is limited to the strength of the input studies and underlying data. As all included studies are observational studies, randomized controlled trials are needed to confirm whether statin can be beneficial in patients with COVID-19 as COVID-19-specific treatment agent. Additionally, there is a general paucity of literature. While there are thirteen studies reporting on mortality, only two studied ICU patients. Only two studies reported on ICU admissions; only one study reported on mechanical ventilation. We need further research to address whether statin is beneficial in terms of preventing ICU admission or mechanical ventilation, which are surrogate markers of severe COVID-19 infection.

In conclusion, patients administered statins after COVID-19 diagnosis were at lower risk of mortality. Ultimately, all included studies in this analysis were observational and predominantly retrospective in nature. Further prospective investigation is needed to assess whether statins may be an effective therapy in COVID-19 patients. Continued investigation is required to assess statin use among COVID-19 patients with respect to other outcomes, such as ICU admissions and mechanical ventilation.

## Data Availability

N/A

## Appendix 1.

Search Strategies

Database: Ovid MEDLINE(R) and Epub Ahead of Print, In-Process & Other Non-Indexed Citations, Daily and Versions(R) <1946 to December 04, 2020> Search Strategy:

--------------------------------------------------------------------------------

1 (Covid-19 or Covid19).mp. (75787)

2 SARS-CoV-2.mp. (24835)

3 severe acute respiratory syndrome coronavirus 2.mp. (37456) 4 or/1-3 (78351)

5 exp Hydroxymethylglutaryl-CoA Reductase Inhibitors/ (41254)

6 statin*.mp. (44888)

7 exp Atorvastatin/ (6644)

8 (Atorvastatin or Lipitor).mp. (9966)

9 (Cerivastatin or Baycol).mp. (784)

10 exp Fluvastatin/ (1398)

11 (Fluvastatin or Lescol).mp. (2109)

12 exp Lovastatin/ (11204)

13 (Lovastatin or Mevacor or Altoprev).mp. (5988)

14 exp Pravastatin/ (3434)

15 (Pravastatin or Lipostat).mp. (4941)

16 exp Rosuvastatin Calcium/ (2530)

17 (Rosuvastatin or Crestor).mp. (3958)

18 exp Simvastatin/ (7807)

19 (Simvastatin or Zocor).mp. (11151)

20 (Pitavastatin or Livalo).mp. (1001)

21 or/5-20 (67164)

22 4 and 21 (162)

23 limit 22 to (english language and yr=“2019 -Current”) (159)

***************************

Database: Embase Classic+Embase <1947 to 2020 Week 49> Search Strategy:

1 (Covid-19 or Covid19).mp. (67997)

2 SARS-CoV-2.mp. (23756)

3 severe acute respiratory syndrome coronavirus 2.mp. (22209)

4 or/1-3 (74109)

5 exp hydroxymethylglutaryl coenzyme A reductase inhibitor/ (159145)

6 statin*.mp. (78372)

7 exp atorvastatin/ (38132)

8 (Atorvastatin or Lipitor).mp. (38991)

9 exp cerivastatin/ (3864)

10 (Cerivastatin or Baycol).mp. (3922)

11 exp fluindostatin/ (9543)

12 (Fluvastatin or Lescol).mp. (3326)

13 exp mevinolin/ (16014)

14 (Lovastatin or Mevacor or Altoprev).mp. (5973)

15 exp pravastatin/ (20066)

16 (Pravastatin or Lipostat).mp. (20522)

17 exp rosuvastatin/ (15501)

18 (Rosuvastatin or Crestor).mp. (15827)

19 exp simvastatin/ (38189)

20 (Simvastatin or Zocor).mp. (39664)

21 exp pitavastatin/ (3255)

22 (Pitavastatin or Livalo).mp. (3341)

23 or/5-22 (180346)

24 4 and 23 (315)

25 limit 24 to (english language and yr=“2019 -Current”) (312)

***************************

Database: EBM Reviews - Cochrane Central Register of Controlled Trials <November 2020> Search Strategy:

--------------------------------------------------------------------------------

1 (Covid-19 or Covid19).mp. (3412)

2 SARS-CoV-2.mp. (1278)

3 severe acute respiratory syndrome coronavirus 2.mp. (246) 4 or/1-3 (3473)

5 exp Hydroxymethylglutaryl-CoA Reductase Inhibitors/ or hydroxymethylglutaryl coenzyme A reductase inhibitor*.mp. (7316)

6 statin*.mp. (10414)

7 exp Atorvastatin/ (0)

8 (Atorvastatin or Lipitor).mp. (5556)

9 (Cerivastatin or Baycol).mp. (172)

10 exp Fluvastatin/ or fluindostatin.mp. (171)

11 (Fluvastatin or Lescol).mp. (722)

12 exp Lovastatin/ or mevinolin.mp. (2263)

13 (Lovastatin or Mevacor or Altoprev).mp. (936)

14 exp Pravastatin/ (1012)

15 (Pravastatin or Lipostat).mp. (2012)

16 exp Rosuvastatin Calcium/ (0)

17 (Rosuvastatin or Crestor).mp. (2605)

18 exp Simvastatin/ (1783)

19 (Simvastatin or Zocor).mp. (3964)

20 (Pitavastatin or Livalo).mp. (532)

21 or/5-20 (19081)

22 4 and 21 (25)

23 limit 22 to (english language and yr=“2019 -Current”) (3)

**************************

## Appendix 2.

PRISMA Flow Diagram

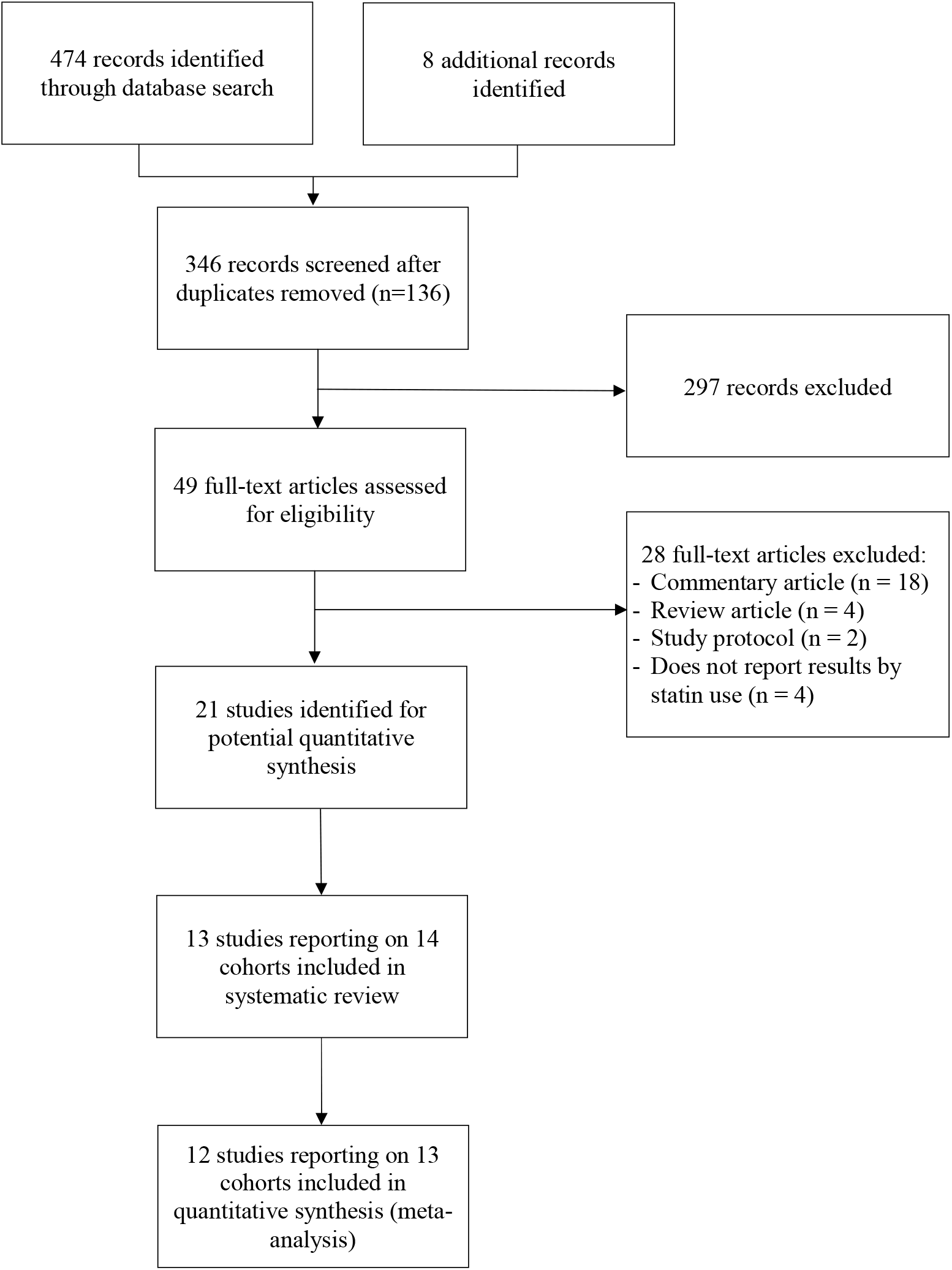

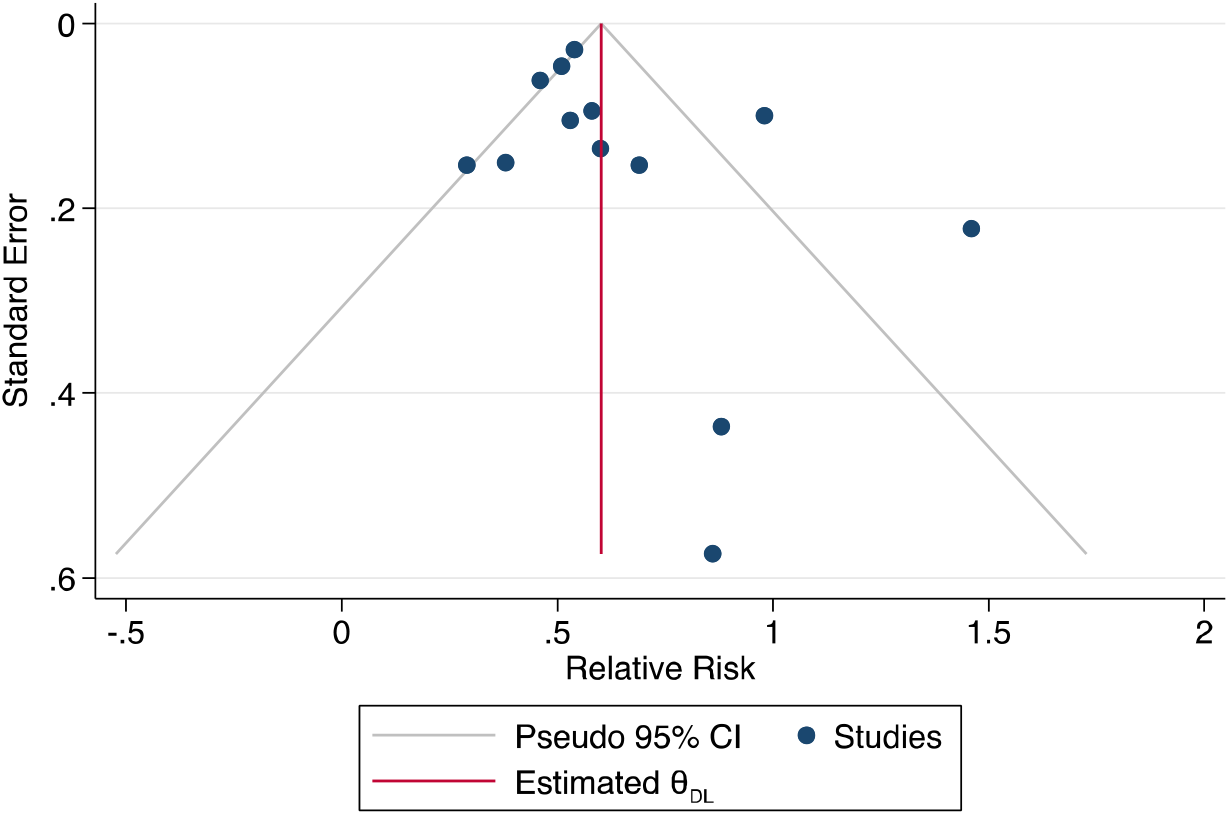

